# The PUPPY Study – Protocol for a Longitudinal Mixed Methods Study Exploring Problems Coordinating and Accessing Primary Care for Attached and Unattached Patients Exacerbated During the COVID-19 Pandemic Year

**DOI:** 10.1101/2021.04.09.21255161

**Authors:** Emily Gard Marshall, Mylaine Breton, Benoît Cossette, Jennifer E Isenor, Maria Mathews, Caitlyn Ayn, Mélanie Ann Smithman, David Stock, Eliot Frymire, Lynn Edwards, Michael Green

## Abstract

**Background:** The COVID-19 pandemic significantly disrupted primary care in Canada, with many walk-in clinics and family practices initially closing or being perceived as inaccessible, pharmacies remaining open with restrictions on patient interactions, rapid uptake of virtual care, and reduced referrals for lab tests, diagnostics, and specialist care. The PUPPY Study (Problems Coordinating and Accessing Primary Care for Attached and Unattached Patients Exacerbated During the COVID-19 Pandemic Year) seeks to understand the impact of COVID-19 across the quadruple aim of primary care, with particular focus on the impacts on patients without attachment to a regular provider and those with chronic health conditions.

**Objective:** The PUPPY Study objective is to understand the impact of COVID-19 across the quadruple aim of primary care.

**Methods:** The PUPPY study builds on an existing research program exploring patients’ access and attachment to primary care, pivoted to adapt to the emerging COVID-19 context. We will undertake a longitudinal mixed methods study to understand critical gaps in primary care access and coordination, comparing data pre- and post-pandemic in three Canadian provinces (Quebec, Ontario, and Nova Scotia). Multiple data sources will be used including: a policy review; qualitative interviews with primary care policymakers, providers (i.e., family physicians, nurse practitioners, and pharmacists), and patients (N=120); and medication prescribing and healthcare billings. The findings will inform the strengthening of primary care during and beyond the COVID-19 pandemic.

**Results:** Funding was provided by the Canadian Institutes of Health Research COVID-19 Rapid Funding Opportunity Grant. Ethical approval to conduct this study was granted in Ontario (Queens Health Sciences & Affiliated Teaching Hospitals Research Ethics Board, file number 6028052; Western University Health Sciences Research Ethics Board, project 116591; University of Toronto Health Sciences Research Ethics Board, protocol number 40335), Québec (Centre intégré universitaire de santé et de services sociaux de l’Estrie, project number 2020-3446) and Nova Scotia (Nova Scotia Health Research Ethics Board, file number 1024979).

**Conclusions:** This is the first study of its kind exploring the impacts of COVID-19 on primary care systems, with particular focus on the issues of patient’s attachment and access to primary care. Through a multi-stakeholder, cross-jurisdictional approach, the PUPPY Study will generate findings and implications for future policy and practice.

## Introduction

### Pre-COVID challenges in Canadian primary care

More than 75% of healthcare visits in Canada are within primary care.[1] Access to primary care is the foundation of a strong healthcare system; vital to achieving the quadruple aim of enhancing patient experience, promoting care-team wellbeing, improving population health, and optimizing costs by managing health in primary care through the life course and reducing burden in acute care.[2] Primary care includes comprehensive and routine care, health promotion, disease prevention, diagnosis and treatment of illness and injury, coordination of care with other specialists and other care services.

Even prior to COVID-19, Canadians reported lower access to a source of regular primary care compared to other Commonwealth nations, with only 90% indicating a regular physician and/or place to receive care in 2020.[3] Access to a regular source of care in Canada, traditionally with family physicians or nurse practitioners, has declined in recent years and varies widely across provinces.[3,4] Individuals without a regular primary care provider are classified as “unattached patients”[5,6] and typically experience poorer health outcomes and less appropriate care than patients with access to a regular primary care provider (i.e., attached patients).[7,8] Vulnerable patients and those with complex needs, including those with low-income levels and/or low social support, are less likely to be attached to a primary care provider, despite benefitting more from access to comprehensive and continuous primary care than less vulnerable patients.[7,9,10] Unattached patients are less likely to seek needed care and often use alternative points of access, such as walk-in clinics, more frequently than attached patients.[11]

As Canadian provinces struggle to support patient attachment to primary care, specific types of care may be provided by community-based pharmacists in some jurisdictions.[12,13] However, primary care provided by pharmacists may not be sufficient and recommended for all patients, particularly those with chronic or complex health concerns and those with needs outside of pharmacists’ legislated scope. Because of these challenges in accessing and being attached to a regular primary care provider, many Canadians rely on emergency departments or walk-in clinics to receive care. Among Canadians surveyed, 42% reported that they had visited an emergency department within the previous 2 years, and among these respondents, 40% indicated their concern could have been treated by a regular primary care provider.[3] Because having a regular primary care provider has been shown to reduce the likelihood of emergency department use,[14] promoting patient attachment to primary care was a key priority prior to the onset of the COVID-19 pandemic. Most Canadian provinces have therefore developed strategies including centralized waitlists for unattached patients and dedicated clinics to address this concern.[15,16]

### Early findings regarding impacts of COVID-19

COVID-19 caused unprecedented disruption to primary care in Canada and internationally. During the peak of the COVID-19 first wave in Canada, many primary care clinics reduced hours,[17] leaving patients and caregivers fearful and uncertain about how to access care. Primary care providers were required to make rapid shifts in practice to comply with infection prevention and control requirements, incorporate COVID-19 triage and non-acute management, address reduced referral and diagnostics access, and implement virtual care where possible.[18–21] Primary care providers had to engage in practice redesign, secure access to personal protective equipment and, in the case of pharmacists in some jurisdictions, integrate changes in scope of practice[22]. Many primary care providers were also redeployed or prepared to be redeployed to COVID-19 testing and treatment roles.[21,23]

Healthcare access is defined as “the opportunity to have healthcare needs fulfilled”[24] and is influenced by a) accessibility of providers, organizations, institutions and systems; and b) the ability of individuals, households, communities, and populations to access primary care. These influential elements have had a COVID-19 “anvil” dropped on their capacity to provide, and access, primary care (see Figure 1).

**Figure 1.**
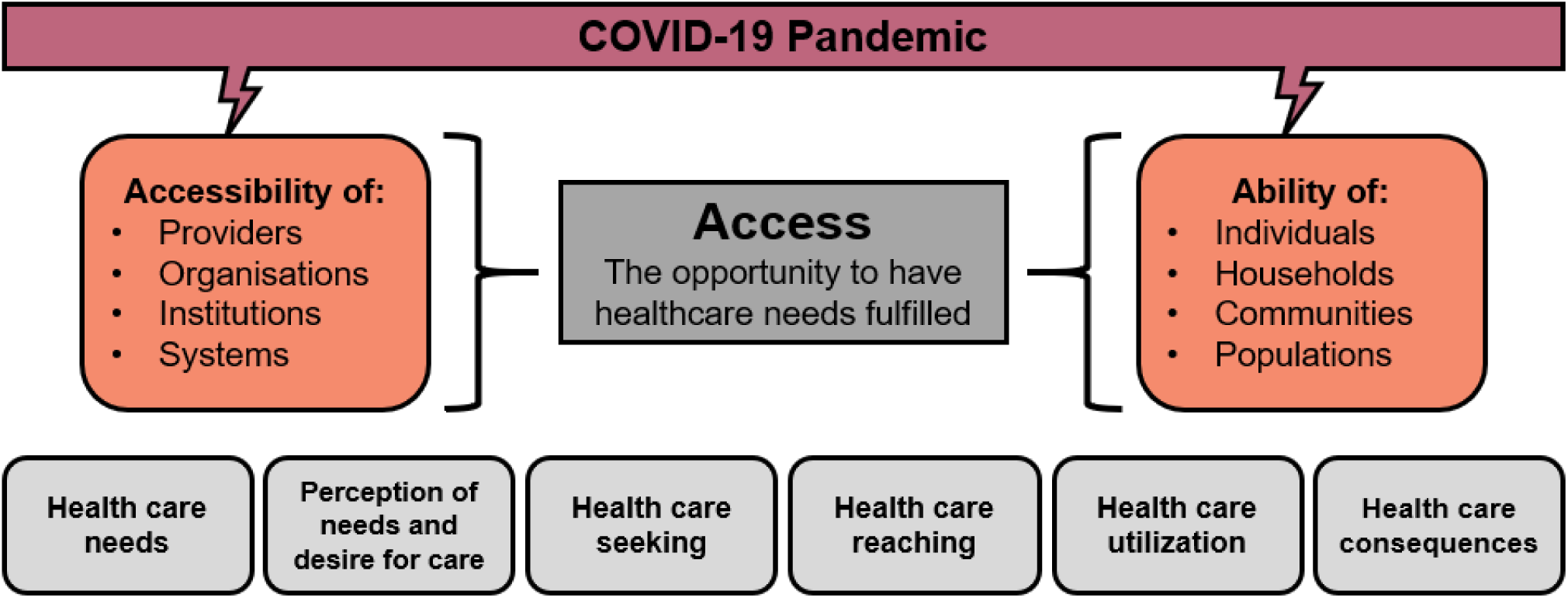
Depiction of the COVID-19-induced disruption to the accessibility of the healthcare system and the ability of patients to access the system (adapted from Levesque et al. 2013)

COVID-19 caused delayed and forgone care concurrent with increased mental health needs of providers and patients. As the pandemic continues, there are anticipated waves of COVID-19 fallout (Figure 2).[25,26] While emerging evidence illustrates some significant impacts of the pandemic on primary care systems globally,[17] the impacts of COVID-19 on patient attachment and access to primary care remain unclear. Evidence is also mounting on the impact of COVID19 on patient and provider wellbeing.[3]

**Figure 2:**
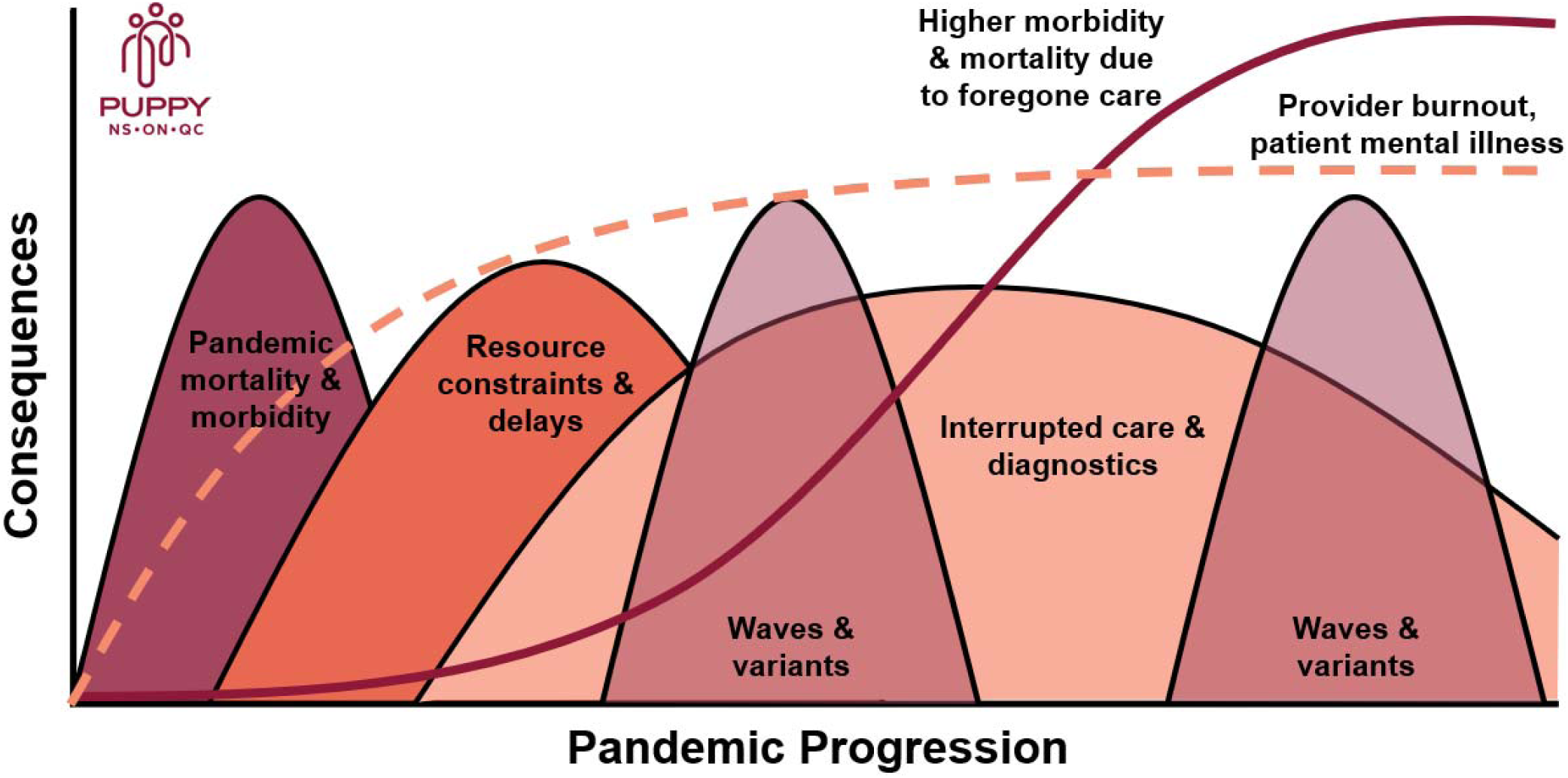
Anticipated waves of COVID-19 pandemic on primary care

### Pivoting a program of primary care research to address COVID-19

At the onset of the COVID-19 pandemic, many health research studies in Canada were required to halt immediately while a pandemic plan and appropriate public health measures were created and enacted. For example, our cross-provincial Canadian Institutes of Health Research (CIHR)-funded CUP Study (Comparative analysis of centralized waitlist effectiveness, policies, and innovations for Connecting Unattached Patients to primary care providers) examining pre-COVID attachment to primary care in three provinces, namely Ontario (ON), Québec (QC), and Nova Scotia (NS) was put on hold for several months.[27]

As the pandemic continued, our team recognized that the existing research aims and methods would not be sufficient to address the impacts of the novel COVD-19 context. Further, new research questions were emerging rapidly due to changes in the policy landscape and provider roles in primary care systems across Canada. For these reasons, it was necessary to pivot existing studies to include pandemic-specific analyses and capture changes in primary care systems over time, while finding novel ways collect data in a safe way during a pandemic. We, the research team, rapidly engaged with our study team, which included Departments and Ministries of Health, Health Authorities, primary care providers and their organizations, and our patient partners. Through these consultations in March and April 2020, we quickly gathered lists of key concerns and priority areas, synthesized and thematically grouped them. The Co-Principal Investigators (EGM, MB, MG, JEI, MM, BC) then developed new strategies for answering emerging questions and updated methods to reflect the new COVID-19 primary health care landscape and ability to work safely. This newly expanded and updated protocol was then submitted for funding in May 2020.

### Objectives

This study will identify and evaluate strategies to provide primary care access and COVID-19 triage and care by family physicians, nurse practitioners, and pharmacists that can be scaled-up to promote attachment and improved access for patients across and beyond COVID-19 waves. We will focus particularly on patients who are unattached, with complex care needs, and/or experiencing social barriers to care, as primary care -based support for these populations may lead to better outcomes for these patients and the healthcare system across the quadruple aim. Objectives include the following:

1. Identify primary care policies and interventions implemented in response to COVID-19 and describe how they affect primary care attachment (i.e., demand) and accessibility (i.e., supply).
2. Understand how COVID-19 related changes affect: i) patients’ experience of accessing primary care, considering different needs, identity factors (e.g., age, gender) and access abilities (un- and attached patients, and/or patients with complex needs); and ii) provider health and wellbeing.
3. Determine how these pandemic-related changes have impacted healthcare utilization, attachment to primary care providers, and medication prescribing, as indicators of access to primary care.
  a. We hypothesize that unattached patients and those with chronic conditions are vulnerable to poorer primary care access and health outcomes exacerbated during the COVID-19 pandemic.
4. Share promising strategies to provide access to primary care with policymakers, primary care providers and patients across Canada in the immediate, intermediate, and reflective phases of the pandemic.

## Methods

### Study design and setting

To address rapid COVID-19 impacts on policy, practice, and patient access to primary care in line with the objectives described above, a longitudinal mixed-methods observational study building from our team’s ongoing research is being conducted. Data will be collected and compared across three cases (NS, QC, and ON) using four methods (see Figure 3) with integration. Data collection will include a) a content analysis of policies impacting primary care access in the wake of COVID-19; b) qualitative interviews with providers, patients, and policymakers; c) surveys of providers and patients; and d) analysis of administrative data, including centralized waitlists, billing and prescribing data, to track healthcare access and utilization, and primary care provider prescribing patterns before, during, and after the COVID-19 pandemic.

**Figure 3.**
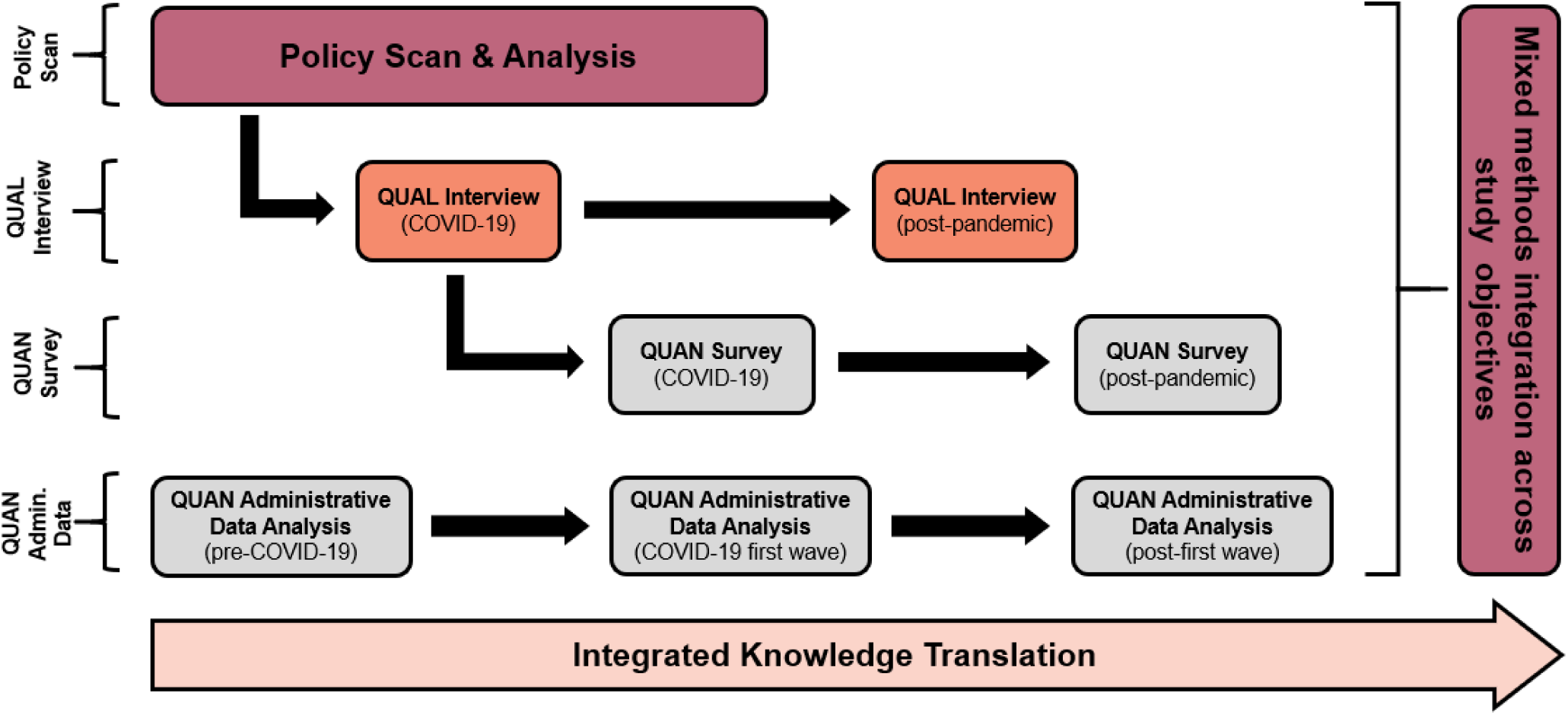
Overview of study objectives, methods, and relationships between activities

### Study participants

A Purposeful sampling approach will be applied to include participant representation from key stakeholders in primary care access including policymakers, patients and primary care providers via qualitative interviews, surveys, and linked administrative health data (see Table 1). While providers from many professions contribute to primary care across systems, our study will focus on family physicians, nurse practitioners, and community pharmacists. The inclusion of the latter is due to the growing number of publicly-funded services offered by pharmacists in several Canadian jurisdictions in recent years, with limited evaluation (e.g., prescribing for minor ailments, immunizations, reviewing and managing medications, etc.),[28–31] which establish more primary care access options.

**Table 1:** Participant groups involved in each of the data collection methods

|  | Data Collection Method |  |  |  | Knowledge Translation |
| --- | --- | --- | --- | --- | --- |
|  | Qualitative | Quantitative | Policy | Administrative |  |

|  | Interviews | Surveys | Content Analysis | Data |  |
| --- | --- | --- | --- | --- | --- |
| **Participant Group** |  |  |  |  |  |
| Patients | ✓ | ✓ |  | ✓ | ✓ |
| Providers | ✓ | ✓ |  | ✓ | ✓ |
| Policymakers | ✓ |  | ✓ |  | ✓ |

### Policy Content Analysis

Contextual factors impacting primary care access will be identified through extensive provincial policy reviews and interviews with health authority, government, regulator, and corporate policymakers. The unit of analysis is provincial. We will document primary care changes in context that coincide with key developments related to COVID-19 to inform recommendations for transformation, scale, and spread. Provincial policies may include: provider hiring and funding, delivery models, including the rapid deployment of virtual care modalities across Canada, incentives, programs and innovations to help patient access, meet the needs of unattached and other vulnerable patients, and other policies that may play moderating roles in primary care (e.g., provider wellbeing). We will focus on influential policies, where policies are defined per the World Health Organization as “decisions, plans, and actions…undertaken to achieve specific healthcare goals” and identified contextual factors.[32]

The Tomoaia-Cotisel approach[33] for assessing and reporting contextual factors of primary care innovations will be applied to the qualitative and policy content analysis components of the study. The framework involves: engaging diverse perspectives, considering multiple policy and context levels, time, formal and informal system/culture, and identifying interactions between policies and contexts. It is tailored specifically to innovations in primary care and considers moderators at multiple levels.

### Qualitative Interviews

Qualitative methods are designed to elicit experiences and perceptions of phenomena where little is known-an ideal approach to study the impact of COVID-19 in primary care. Stakeholder groups to be interviewed include patients, family physicians who do and do not take on new patients, nurse practitioners, community pharmacists, and policymakers with roles relevant to primary care access and attachment. The proposed longitudinal data collection will support interviews, n=10 participants per stakeholder group per province (N=120 participants in total), which will ensure saturation.[34,35]

Interviews will be conducted during COVID-19 and post-pandemic to elicit current and retrospective lived experiences. Interview guides will be developed to reflect key issues pertinent to stakeholders. For example, providers will be asked questions pertaining to practice changes. Patients will be asked about their experience with primary care changes and the impact of these on access and wellbeing. Policymakers will be invited to share processes for, and outcomes of, policy change and will be consulted on relevant documents to include in our policy content analysis for Objective 1.

Purposive and snowball sampling strategies will be used, stratifying by relevant participant characteristics (gender, rurality, practice characteristics, etc.). Invitations for interview participants will be distributed via the provincial centralized waitlists, partnered organizations and social media. We will iteratively revise our sampling and recruitment strategies as we collect data and learn more about patient and provider experiences.[36]

Informed consent discussions and semi-structured in-depth interviews will be conducted virtually using Zoom Videoconferencing (Zoom Video Communications Inc.) by a Masters-trained researcher. Audio recordings of interviews will be transcribed verbatim and coded in NVivo software (QSR International). Coding reports will be generated and examined to uncover themes and patterns in the data.

Preliminary thematic analysis will provide rapid reporting to stakeholders. A Framework Analysis[37] approach will incorporate the Levesque et al. conceptual framework[24] for access to healthcare and be implemented across study phases for comparative analysis. This method allows for inductive and deductive coding approaches.[37] We will code deductively to the Levesque framework and inductively from interview transcripts, allowing emergent themes to enhance what can be gleaned from the framework alone. Intra and cross-case analysis will be conducted by incorporating provincial framework analysis matrices.[37,38]

### Surveys

Brief surveys for patients and providers will be developed to determine prevalence of our emerging qualitative themes. Surveys designed for providers will be delivered via the secure online Opinio Survey Tool (ObjectPlanet, Inc.) post-pandemic to measure the degree to which COVID-related policy changes have impacted primary care access and attachment, as well as their personal wellness. Recruitment support will be provided by our partners. An online patient survey at the same time point will explore patient primary care access and attachment during COVID-19. A convenience sample of 1000 patient respondents per province will be recruited using a third-party survey sampling company (N=3000). It is estimated that a sample size of 1000 per province would permit adequate segment sizes for comparison of results among patient groups and provinces. The use of third-party sampling services are common for healthcare research involving the general public.41-43 Bivariable and multiple regression models will be generated to show trends and associations on key elements across phases. Follow-up surveys will be conducted at a later point to assess changes over time.

### Administrative Data

Analyses of pre-COVID-19 prescription dispensation, centralized waitlist, physician billing, and inpatient and outpatient hospital discharge data has already begun to examine effectiveness of centralized waitlists for a related study.[27] As part of the PUPPY Study, we will expand this analysis to explore changes across pre-COVID-19, COVID-19, and post-pandemic periods. Harmonized indicators of health care utilization (e.g., primary care, emergency, hospitalization, and potentially avoidable inpatient care), and primary care attachment indicators (primary care provider attachment, continuity of primary care), and primary care service provision (e.g., frequency and type of primary care encounters, continuity of medication dispensation for maintenance of chronic conditions) will be measured across the three participating provinces. Change in these indicators, and in care continuity, will be estimated and compared over pandemic wave-indexed study periods.

Multivariable regression will be used to identify potential clinical (e.g., patient complexity, comorbidity) demographic and socioeconomic determinants of primary care need, and changes in these indicators over the course of the pandemic. Socioeconomic determinants are derived from the 2016 Canadian census data, including the Canadian Index of Multiple Deprivation, with a focus on dimensions of economic dependency, ethno-cultural composition, and situational vulnerability.[27] Centralized waitlist data will be used to measure primary care attachment and assess changes in access to primary care. Building on ongoing work, variation in patient primary care provider attachment rates, demand for attachment, and time to attachment among those identified on centralized wait lists will be quantified and changes in these outcomes assessed across study periods. Determinants of these outcomes will be identified, and their relative magnitude estimated, using multivariable techniques. In each province, study populations will be stratified by age, sex (and gender where feasible), degree of comorbidity and geography (i.e., urban versus rural) to identify those at greatest risk of being unattached to a primary care provider.

### Mixed Methods Integration

As a longitudinal study comparing three provincial cases, PUPPY will employ a series of mixed methods integration approaches and principles to inform the planning, analysis, and interpretation across the four data types.[39] Adapted from Goldsmith et al.[40], Figure 4 provides a depiction of the ways in which the four study methods will inform one another and ultimately lead to meta-inferences strengthened by this mixed methods approach.

**Figure 4:**
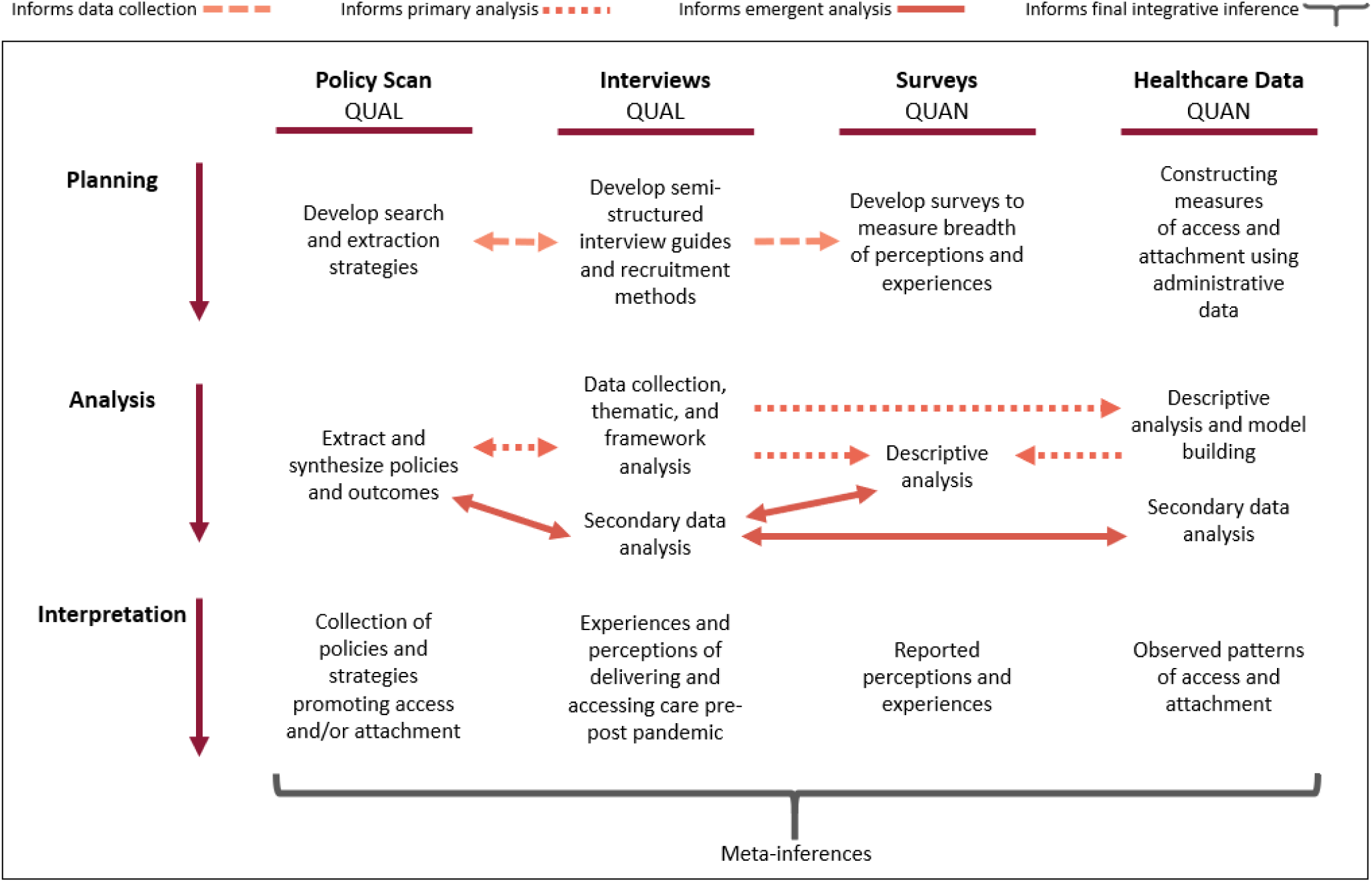
Summary of mixed methods integration approaches across the planning, analysis, and interpretation stages of the PUPPY Study

In the planning phase, qualitative work and policy content analysis approaches will be conducted in parallel, with findings from each iteratively informing data collection and planning for the other. For example, qualitative interviews will support identification of policy documentation unable to be identified through traditional searches, while analysis of policy documents will uncover areas of interest to explore in future qualitative interviews. Additionally, in an exploratory sequential approach, qualitative interview findings will be used to inform development of a quantitative survey to build upon and explore the breadth and depth of perceptions expressed by interview participants.

In the analysis phase, data will be brought together through embedding and merging, a process by which multiple datasets are brought together for analysis and triangulation via iterative comparison.[39] This process will enable creation of rich case descriptions. In particular, a timeline for each case (province) illustrating the patterns in healthcare data alongside policy milestones and insight into relevant participant experiences will be developed. The frameworks being used to inform our approaches have been used across multiple methodologies and mixed methods designs and will facilitate these comparisons.[24,33] As seen in Figure 4, use of congruent methods will allow numerous comparisons between datasets for both primary and emergent research questions. Mixed methods interpretation will be conducted via the creation of mixed methods narratives and joint displays from which meta-inferences incorporating multiple methodologies can be generated.[39]

## Results

### Funding

In June 2020, our team received funding through the CIHR COVID-19 Rapid Funding. The funding opportunity encouraged an expansion of ongoing studies to expedite translation of findings and offered resources to identify and incorporate emerging research questions, expand existing methods, and include additional methods where necessary. Through this opportunity, our team received the resources necessary to elucidate the impacts of COVID-19 on primary care in Canada via the current study.

### Ethics

Approval to conduct this study was granted in Ontario (Queens Health Sciences & Affiliated Teaching Hospitals Research Ethics Board, file number 6028052; Western University Health Sciences Research Ethics Board, project 116591; University of Toronto Health Sciences Research Ethics Board, protocol number 40335), Québec (Centre intégré universitaire de santé et de services sociaux de l’Estrie, project number 2020-3446) and Nova Scotia (Nova Scotia Health Research Ethics Board, file number 1024979). As the PUPPY Study builds upon and expands the timeline of ongoing research projects, including the CUP Study,[27] ethical approvals have in several cases been granted as amendments or extensions to the CUP Study to facilitate rapid implementation of study activities.

### Timeline

Data collection for the PUPPY study will take place in 2021-2022, with rapid reporting taking place between 2021-2023. As of April 2021, recruitment for qualitative interviews has begun in NS and QC, with recruitment expected to take place in ON when COVID-related pressures have eased. Each province is in the process of accessing administrative health data and linking it to provincial centralized waitlist data. Integrated and end-of-grant knowledge translation of the PUPPY Study and subsequent research will follow up on key areas identified.

## Discussion

### Partnership and Knowledge Translation

Our team includes regulatory bodies and associations representing family physicians, nurse practitioners, and community pharmacists, as well as support from the Canadian Institutes of Health Research Strategy for Patient Oriented Research (SPOR) Primary and Integrated Healthcare Innovations (PIHCI) Networks and the SPOR Support for Patient-Oriented Research and Trials Units to aid data collection and knowledge dissemination. Guidance from COVID-19 policymaking partners will ensure relevance and uptake while minimizing burden of study activities on participants, which is particularly critical given the high demands of the pandemic on all stakeholders involved in our study. Data collection activities will occur remotely to comply with public health measures. In anticipation of possible participant distress, and recognizing the impact of the pandemic on mental health and wellness generally, we will provide a list of resources to appropriately trained mental health and primary care providers.

To ensure appropriate dissemination and translation of study findings, all data collection begins with consultation. Team members representing all stakeholder groups, including providers, policymakers, and patients, will participate in development and refinement of study tools, analysis, interpretation, and dissemination plans. Knowledge dissemination will include multiple modalities to maximize uptake of findings. Policy briefs and reports will be shared at each study phase and assisted by professional graphic and communication design support. Other modalities include peer-reviewed publications, conference presentations, local presentations to key stakeholder groups (e.g., provider associations, health authorities, Departments/Ministries of Health, primary care provincial leadership meetings), knowledge sharing on departmental websites, blog posts, and social media. Team members, including patient partners, will have the opportunity to inform, author, and participate in dissemination activities. Using CUP study funds post-pandemic, we will facilitate cross-jurisdictional learning via a symposium with stakeholders from across Canada to improve primary care attachment and manage patients within and outside of pandemics.

### Conclusion

The PUPPY study is designed to provide rapid support for primary care policymaking, provider needs, and patient access to primary care from investigation across the COVID-19 waves. We will regularly communicate emerging recommendations to our partners for timely policy optimization. Immediate term early data collection will provide feedback on new policies in primary care settings and impacts on patient access, providing insight into possible unintended consequences of rapid policy transformation and revealing promising strategies. This information will inform provision of care through changing pandemic contexts, including requirements for physical distancing and safety requirements. In the intermediate term, our study will document changes in the primary care policy landscape to strengthen the response to additional “waves” related to COVID-19. Findings will be distributed to study partners and beyond via our networks, (e.g., CanCOVID, pan-Canadian PIHCI Networks, North American Primary Care Research Group), to support cross-jurisdictional pandemic response. In the long term, findings will help us grasp the impact of these policy changes and events on the ability of systems and providers to coordinate and deliver primary care, patient access to primary care, and on health outcomes. Recommended best practices to improve access to primary care as we transition to a post-pandemic context will be widely shared with our partners via our knowledge dissemination plan as outlined above.

## Data Availability

No data associated with this study are available at time of development of this protocol.

## Acknowledgements

We would like to acknowledge the generous support of our PUPPY study organizational partners and team members, including our academic researchers, providers, policymakers, and patient partners Sarah Peddle, Ana Correa Woodrow, Nicole Desjardins, and Danièle Roberge. A full list of our team members and partners can be found elsewhere.[41] We would like to thank the Canadian Institutes of Health Research for their support in the form of a COVID-19 Rapid Funding Opportunity Grant.

## Abbreviations

PUPPY: Problems Coordinating and Accessing Primary Care for Attached and Unattached Patients Exacerbated During the COVID-19 Pandemic Year (current study title)
CIHR: Canadian Institutes of Health Research
NS: Nova Scotia (Canadian province)
ON: Ontario (Canadian province)
QC: Québec (Canadian province)
CUP: Comparative analysis of centralized waitlist effectiveness, policies, and innovations for connecting unattached patients to primary care providers (study title)

